# The effect of short-graft preparation with tape suspension and screw fixation on loss of knee extension following anterior cruciate ligament reconstruction: A retrospective cross-sectional analysis of public hospital cases from 2015 - 2017

**DOI:** 10.1101/2020.04.21.20073494

**Authors:** Christopher Bell, Corey Scholes, Maha Jegatheesan, Kirby Tuckerman

## Abstract

**Introduction:** The short graft with tape suspension (SGTS) is a technique for ACL reconstruction that has gained popularity in recent years. Though the construct utilises a hamstring tendon, its biomechanical properties more closely resemble a stiffer graft such as bone-patella-bone. Due to these properties, there are concerns this technique may increase the likelihood of postoperative loss of extension (LOE), particularly if the surgeon does not modify their tensioning technique. This study compared LOE in patients undergoing ACLR with the SGTS technique, versus other ACLR techniques. We hypothesised that with appropriate technique modifications, the SGTS technique would not be inferior to long hamstring graft techniques with respect to LOE observed clinically during supervised rehabilitation.

**Materials and Methods:** We retrospectively reviewed 138 patients who received primary ACLR at one of two hospitals between January 2015 and December 2017 and elected to participate in a rehabilitation program with the hospital physiotherapy department. Postoperative knee extension was assessed by a department physiotherapist until satisfactory function was achieved. Patients were classified as SGTS ACLR or non-SGTS ACLR during chart review and LOE compared at initial assessment and at the time of maximum extension, via a noninferiority analysis.

**Results:** The grafts for the SGTS group (N=44) were significantly larger in diameter (median 8.5mm vs. 8.0mm, P <0.001) and less incidence of notchplasties (17.8% vs. 44.7%, P <0.001) compared with the non-SGTS group (N=94). The upper 95% confidence interval for the difference in proportions between groups did not exceed the non-inferiority margin (0.3 or 30%) at either Initial or Maximum timepoints.

**Conclusions:** The SGTS technique was not inferior to other hamstring-graft ACLR techniques with respect to postoperative LOE. Surgeons using or considering using the SGTS construct can rule out increased incidence of LOE as a factor in their decision-making, providing the grafts are prepared according to existing guidance and tensioned in full extension. Further studies are recommended to assess longer term functional outcomes and ultimately treatment success.

## 1. Introduction

The goal of anterior cruciate ligament (ACL) reconstruction is to restore the injured knee to its pre-injury biomechanical state [15]. To achieve this, the knee must be both stable and have normal range of motion, particularly full extension [27]. Both surgical and non-surgical factors contribute to these outcomes.

Surgical factors include the amount of tension placed on the graft at the time of reconstructive surgery and the position of the knee when this tension is applied [1]. Applying high levels of tension, particularly with the knee in flexion, may increase stability but at the expense of regaining full extension. This loss of extension (LOE) results in asymmetric limb loading, delays in reaching rehabilitation milestones and higher rerupture rates when the athlete returns to sport [13, 17]. LOE has also been shown to correlate strongly with poorer subjective outcome scores following ACL reconstruction [27].

To achieve the right balance between range of motion and stability, the surgeon must be familiar with the biomechanical properties of the chosen graft. Hamstring (HS) tendon grafts are known to have lower stiffness and higher likelihood to stretch over time than bone-patella tendon-bone (BPTB) grafts [5]. To compensate for this, it is common for surgeons to tension HS grafts with the knee in flexion. In contrast, most surgeons tension BPTB grafts in full extension [16, 20].

Short-graft HS ACL reconstruction has gained popularity over the last decade. One such system is the Tape Locking Screw (TLS®) which has a number of attributes that may place the patient at increased risk of LOE if tensioned with a similar technique to traditional HS grafts. The average graft diameter is larger than long HS graft techniques [22]. The graft is shorter, making elongation less likely as this is a function of both elasticity and initial graft length. Very little slippage has been shown between the tape and screw fixation in comparison to other devices [24]. Finally, the graft is pretensioned with 500N for 1-2 minutes, theoretically minimising further elongation of the graft after implantation [6, 23].

The optimal tensioning technique for short-graft ACL reconstruction has not been well defined. Due to its biomechanical properties, the senior author chose to tension the TLS graft in full extension due to concerns for LOE. To our knowledge, this is the first study to examine the association between short-graft ACL reconstruction and post-operative LOE. We hypothesised that with modification to traditional hamstring ACL reconstruction techniques, the TLS method would be non-inferior to other hamstring graft techniques with respect to incidence of postoperative LOE.

## 2. Materials and methods

### 2.1 Study setting

This was a retrospective dual-centre study of ACLR performed on patients between January 2015 and December 2017 at two metropolitan hospitals offering Government/Public healthcare (Queen Elizabeth II Jubilee Hospital [QEII] and Mater Private Hospital Springfield [MPHS]). All patients had presented to QEII for initial consultation. Patient lists were sourced from a previous dataset [29] with patients confirmed to have participated in rehabilitation at the QEII physiotherapy department, at the time of introduction of the SGTS technique. A second patient list was sourced from the department clinical registry (SHARKS, ACTRN12617001161314) to include patients not in the [29] dataset. Chart review was used to confirm the status of the second patient list against the inclusion/exclusion criteria.

#### Inclusion criteria

- Electing to undergo primary ACL reconstruction at either hospital
- Participation in post-reconstruction rehabilitation at the QEII hospital outpatient physiotherapy department.
- Isolated ACL injury +/- meniscal injury only

#### Exclusion criteria

- Revision ligament reconstruction procedure
- Concurrent surgical treatment of other ligamentous injury (PCL, LCL, MCL, PLC)
- Receiving physiotherapy external to the hospital outpatient department

As per the hospital’s usual practice, new patient referrals were triaged by a consultant orthopaedic surgeon and assigned to a lower limb surgeon. The decision to proceed to surgery was made between the surgeon and patient and was not protocolised. From July 2017, patients booked for surgery were also captured in the department clinical registry (SHARKS) and consented using a written-informed opt-in approach. Ethical approval was granted for the registry by the Metro South Health Human Research Ethics Committee (HREC/16/QPAH/732).

### 2.2 Surgical Technique - Short Graft, Tape Suspension

During the period of interest, 14 surgeons performed ACLR of which two used a SGTS technique. All patients were examined under anaesthesia. Tendon(s) were harvested prior to commencing arthroscopy. The standard TLS® surgical technique was used and has been described in detail by Collette and Cassard [6]. The semitendinous tendon was harvested in isolation and Gracilis was used only in the event of graft failure or insufficient graft material. The graft length was set at 50 - 55mm. Prior to implanting the graft, arthroscopic examination of the knee was undertaken. Any meniscal injuries were addressed using an all-inside technique repair or resected to stable margins. The prepared graft was inserted into the knee via the anterior-medial portal. Once the graft was adequately positioned and fixed into the femur, the graft was then tensioned with a “sardine-can” technique with the knee in full extension before the tibia screw was inserted. The knee was then examined arthroscopically in full extension to evaluate the intercondylar notch and notchplasty was performed if required.

### 2.3 Surgical technique – Non-SGTS

The remaining patients had ACLR by 12 surgeons using non-SGTS techniques, comprising femoral closed-loop button/tibial interference screw fixation, femoral cross-pin/tibial interference screw and femoral and tibial interference screws. Included cases received hamstring (either semitendinosus or semitendinosus and gracilis) autografts from the ipsilateral leg. There were two cases where the hamstring graft was reinforced with lateral tenodesis (iliotibial band). Grafts were fixed with the knee in 20-30° flexion. Meniscal pathology was addressed intraoperatively either by partial meniscectomy or primary repair. Notchplasty was performed as required. Arthroscopic approach and graft placement were as per surgeon preference.

### 2.4 Rehabilitation

Following ACLR, patients at both hospitals were reviewed by the ward physiotherapist with identical post-operative instructions and referred for outpatient physiotherapy at QEII hospital. The aim was for all patients to have an initial assessment within 10 days of surgery. Frequency of appointments was based on patient progress. A four-phase, goal-directed program was used as described in the Randall Cooper ACL Rehabilitation Guide [7]. Patients were discharged from outpatient physiotherapy once they achieved baseline function and were provided with written information regarding ongoing rehabilitation, risk of re-injury and osteoarthritis.

### 2.5 Data collection

A list of all patients who had a primary ACLR at QEII and MPHS between January 2015 and December 2017 was sourced from hospital systems. Patients who had participated in physiotherapy at QEII following surgery were included in the review. Knee ROM was collected and documented at each physiotherapy session using a standard universal goniometer with the patient seated. Operative notes from those patient charts were used for ACLR technique and arthroscopic findings. Physiotherapy examination and progress were also recorded in the patient charts.

#### Primary Outcome Measure

Extension angle (°) measured by the physiotherapist during in-person visits was dichotomized into loss of extension (LOE) (yes; no) using a threshold of >5° compared to the non-operated limb. Shelbourne [31] reported that 5° LOE (relative to contralateral limb) was a strong predictor of low patient-reported scores postoperatively. LOE was extracted from the chart for the first physiotherapist session (Initial) and when maximum extension was measured during the course of rehabilitation prior to formal discharge (Maximum).

### 2.6 Data and Statistical Analysis

Data were retrieved from a sample of convenience, based on clinical volume before and after the introduction of the SGTS technique, changes to electronic medical records processes and data availability. At the conclusion of the chart review, the spreadsheet was locked to further editing and prepared for analysis. Responses were recoded for graft type to standardise into response groups (ST, STG).

#### Missing Data

Missing data were assessed for each variable across both timepoints (initial and maximum), as well as the pattern of missingness using the *misstable* function in Stata. The missingness was limited (0.7 - 2.1%) for the key covariates identified as important in the directed acyclic graph. Extension measurements were missing for a higher proportion of cases than the remaining covariates (6%); these were imputed as *No LOE present*. It was assumed that if a concern regarding extension was present, this would have been noted in the chart. Complete case analysis was used to estimate the adjusted proportion estimates for each SGTS group at Initial and Maximum timepoints, assuming that the missingness occurred completely at random (MCAR). Considering the mechanism of data generated in the charts and the low overall missingness, imputation was not used to estimate missing values.

#### Statistical Analysis

A directed-acyclic-graph (DAG) method was used to map the relationships between the estimand (LOE incidence) and the primary exposure (use of short graft with tape suspension or not) to establish a reasonable model. A DAG is a non-parametric diagrammatic presentation of the assumed data-generating process for a set of variables in a specified context. Variables and their measurements are depicted as nodes (or vertices) connected by unidirectional arcs (or arrows;hence ‘directed’) depicting the hypothesised relationships between them (Tennant et al. 2020). Confounders are identified by an open path (all arrows between nodes point in the same direction) passing through a node between the exposure and the estimand. To reduce confounding bias, the confounders identified in the DAG can be included in a multivariable model as covariates (Tennant et al. 2020). In this study, a DAG was constructed using the browser-based version of Daggity v3.0 (Textor et al. 2016) (**Figure 1**). The following confounders were identified and included in the model;

- Sex (male; female)
- Injury to surgery delay (days)

The following covariates that may improve precision of the estimand were identified and included in the model;

- Age at surgery (years)
- Body mass index (measured at time of surgery booking) (kgm^2^)
- Presence of secondary pathology (meniscal or chondral lesions) (yes; no)

**Figure 1:**
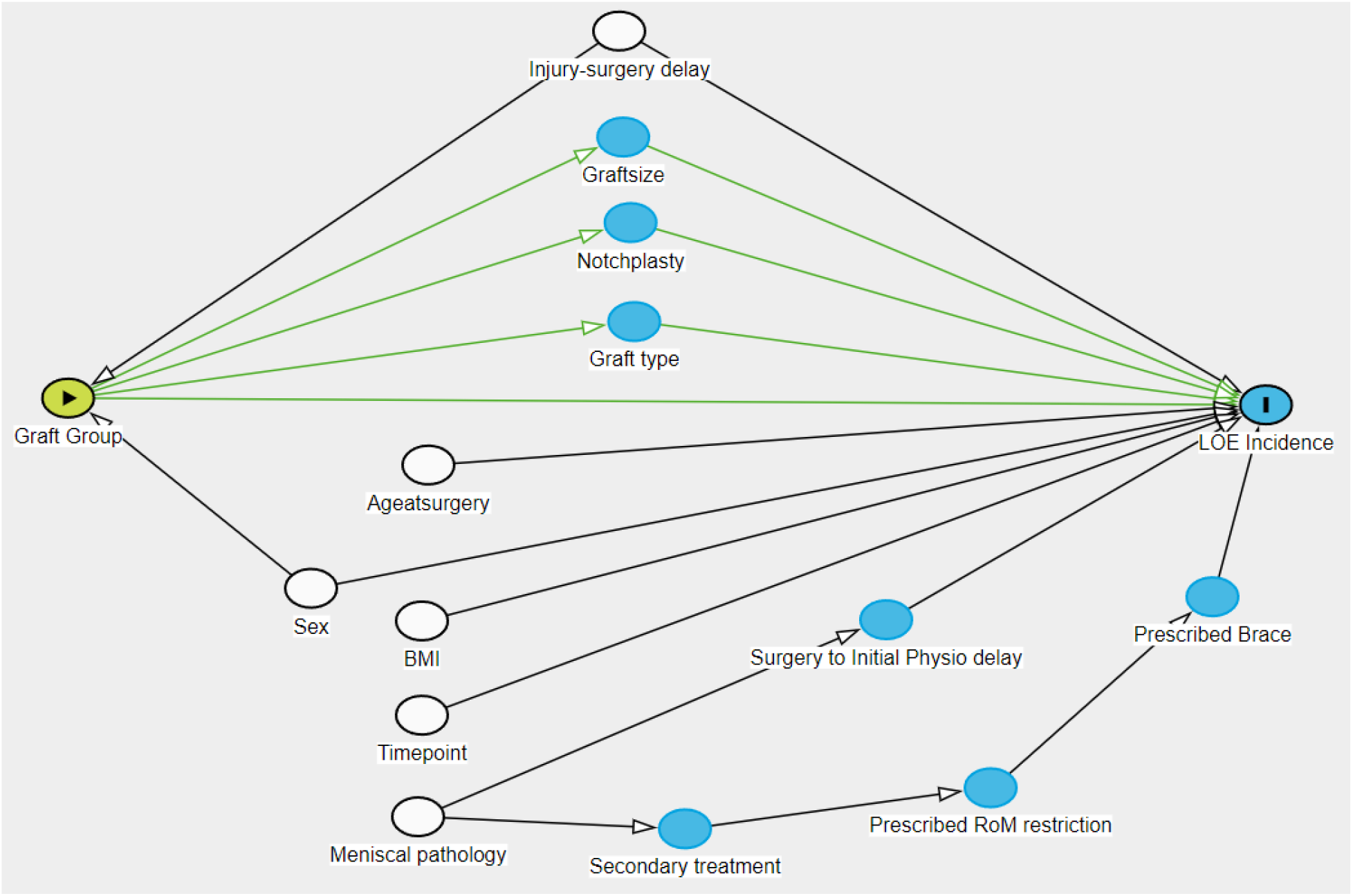
Directed acyclic graph illustrating the potential relationships between *graft group* (exposure) and *LOE incidence* (outcome)

The DAG also identified a number of mediators present in the dataset that should not be included in the model (e.g. Graft Size; **Figure 1**). A mixed-effects binary logistic regression model with *LOE present (yes/no)* as the outcome was used to create adjusted estimates of *LOE incidence* (0 - 1) for each SGTS group at each timepoint (Initial vs Maximum). Patient ID was included as a random effect to accommodate the potential correlation between extension measurements within a patient between timepoints.

The period between surgery and initial physiotherapy appointment was excluded from the model due to its dependence on the presence of secondary pathology and to reduce the risk of model overfitting. The interaction term between *SGTS group* and *timepoint* was tested and the effects of covariates assessed with an alpha of 0.05. Marginal estimates based on model predictions were derived for each SGTS group and *timepoint* combination. Non-inferiority was declared if the upper limit of the one-sided 97.5% confidence interval for the marginal estimate of *LOE Incidence* in the SGTS group did not exceed a relative difference of 30% from the baseline incidence of the *non-SGTS* group (risk-ratio 1.3), equivalent to a one-sided test with an alpha of 0.025 [28]. The non-inferiority margin was derived from a related systematic review [10] which identified a median baseline incidence of LOE 0.16 at a median of 4.9 months post ACLR. A 10% absolute increase in patients presenting with LOE given the new technique above the rate seen with the existing techniques was deemed clinically acceptable. A 10% absolute increase in incidence was converted to a relative increase using the equation AR/BI, where AR is the absolute acceptable margin and BI is the baseline incidence. This gave a margin of 0.6, which was halved to 0.3 (5% absolute increase) to provide a conservative non-inferiority margin. Model and marginal estimates were calculated in Stata (v17.1, StatCorp LLC, USA) using the *melogit* and *margins* commands (see Supplementary material).

## 3. Results

A sample of 138 patients who met the inclusion criteria were included in the final analysis (**Figure 2**). All except 2 patients had an extension measurement recorded at initial postoperative appointment with a median for non-zero cases (proportion 0.69 95%CI 0.60 - 0.76) of 10° (IQR 5 - 10) and a median of 5° (IQR 5 - 7.8) for non-zero maximum extension (proportion 0.26, 95%CI 0.19 - 0.34) recorded prior to termination of the rehabilitation program (patient self-selected, therapist discharge, failed to attend). The sample was majority male, aged in mid-20s, with BMI slightly above the boundary for *normal* (**Table 1**). Missing data rates ranged from 0 - 7%. The mixed-effects model revealed that the covariates included were not significantly associated with LOE incidence in isolation (**Table 2**), although there was a significant reduction in LOE for both groups at *Maximum* measurement compared to Non-SGTS at Initial measurement. In contrast, considerable patient variation was observed as denoted by the relative size of the coefficient for the *Patient* factor (**Table 2**). Predicted estimates for LOE incidence ranged from 0.15 to 0.70 between the two groups at the two timepoints (**Table 3**). The upper 95% confidence interval for the difference in proportions between groups did not exceed the non-inferiority margin (0.3 or 30%) at either Initial or Maximum timepoints (**Figure 3**).

**Figure 2:**
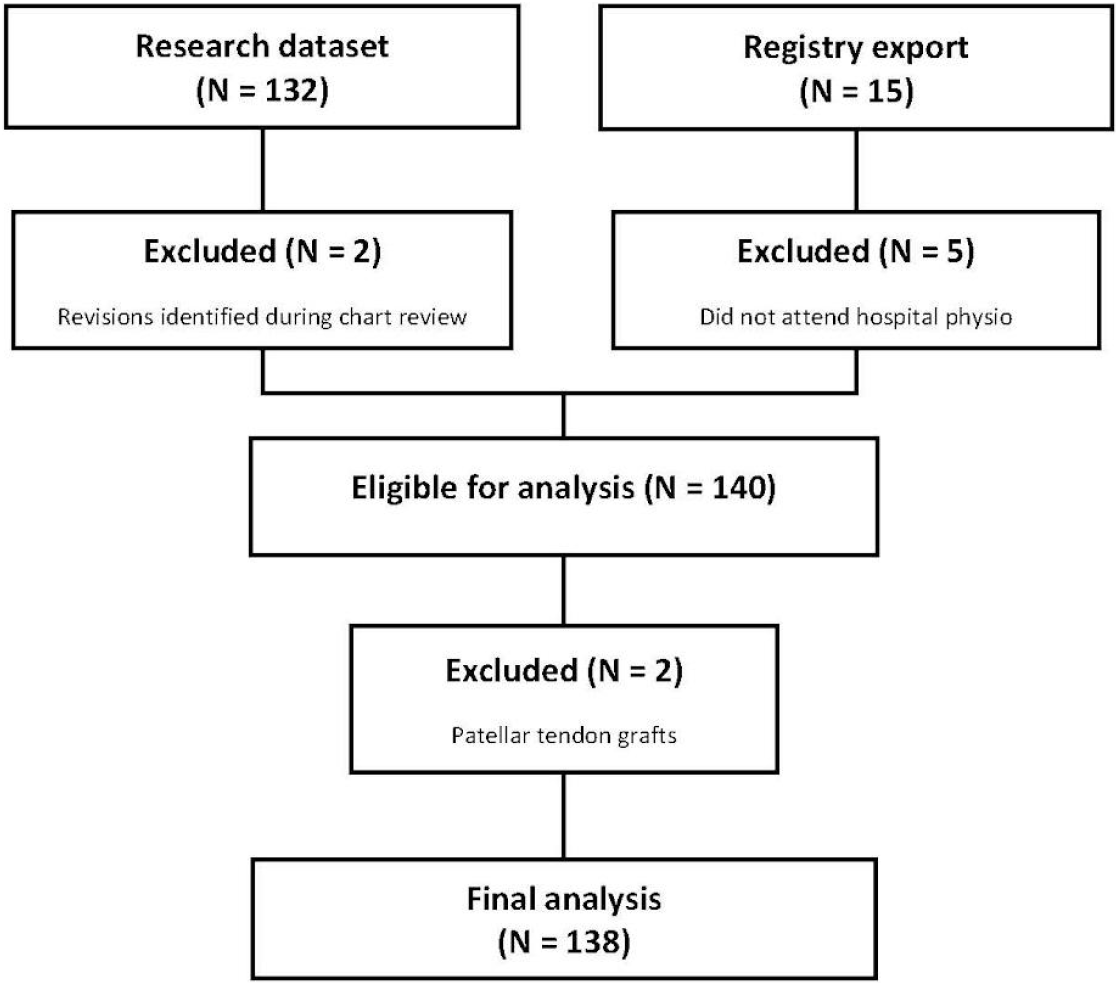
STROBE [30] flow chart of inclusion, exclusion and analyses.

**Figure 3:**
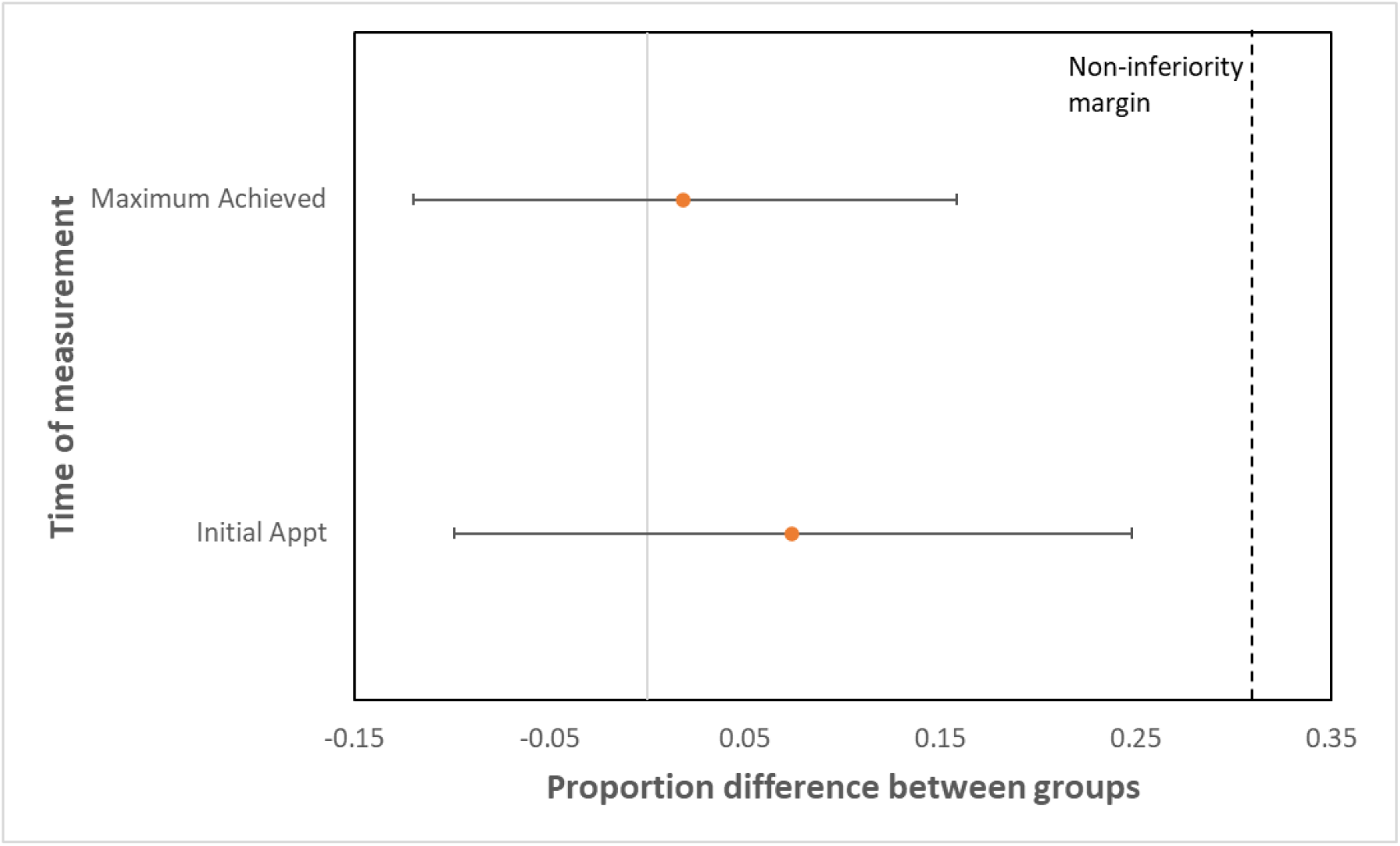
Adjusted differences in proportions with 95% confidence intervals between SGTS vs non-SGTS groups for loss of extension (LOE) at initial physiotherapy appointment and at the time maximum extension was achieved during rehabilitation. Positive difference and positive confidence interval indicates higher incidence of LOE in SGTS group.

**Table 1:** Summary characteristics of patients sampled in analysis (median with interquartile range)

|  | **SGTS** | **Non-SGTS** | **Cohort** | **Missing data (N)** |
| --- | --- | --- | --- | --- |
| N | 45 | 94 | 139 |  |
| Age at Surgery (years) | 25.7 (18.6 - 31.4) | 26.4 (21.6 - 31.4) | 26.1 (21.2 - 31.5) | 0 |
| Male (%) | 46.7 | 70.2 | 62.6 | 0 |
| BMI (kg/m<sup>2</sup>) | 26.1 (24 - 29.1) | 26.3 (23.7 - 28.3) | 26.2 (23.7 - 28.4) | 0 |
| Date of Injury to Date of Surgery (weeks) | 21.6 (13.9 - 38) | 38 (23.6 - 62.3) | 29.6 (18.7 - 58.4) | 8 |
| Graft Type (%)<br>ST<br>STG | 71.1<br>28.9 | 4.3<br>93.6 | 25.9<br>72.7 | 2 |
| Graft size (mm) | 8.5 (8 - 9.5) | 8 (7.5 - 8.5) | 8 (7.5 - 8.5) | 6 |
| Notchplasty (%) | 18.2 | 44.7 | 36.2 | 0 |
| Meniscal pathology (%)<br>- Medial<br>- Lateral | 43.2<br>34.1 | 37.2<br>36.2 | 39.1<br>35.3 | 0 |
| Secondary treatment (%) | 52.3 | 54.3 | 53.6 | 2 |
| Prescribed RoM restriction (%) | 25 | 46.8 | 39.9 | 0 |
| Date of Surgery to Date of Initial Physio Appt (weeks) | 1.8 (1.3 - 2.3) | 1.6 (1.1 - 2.1) | 1.6 (1.1 - 2.1) | 2 |
| Initial Physio Appt to Maximum Extension (weeks) | 6.4 (1.2 - 14.6) | 2.6 (0 - 6.7) | 3 (0 - 8.1) | 2 |

**Table 2:**
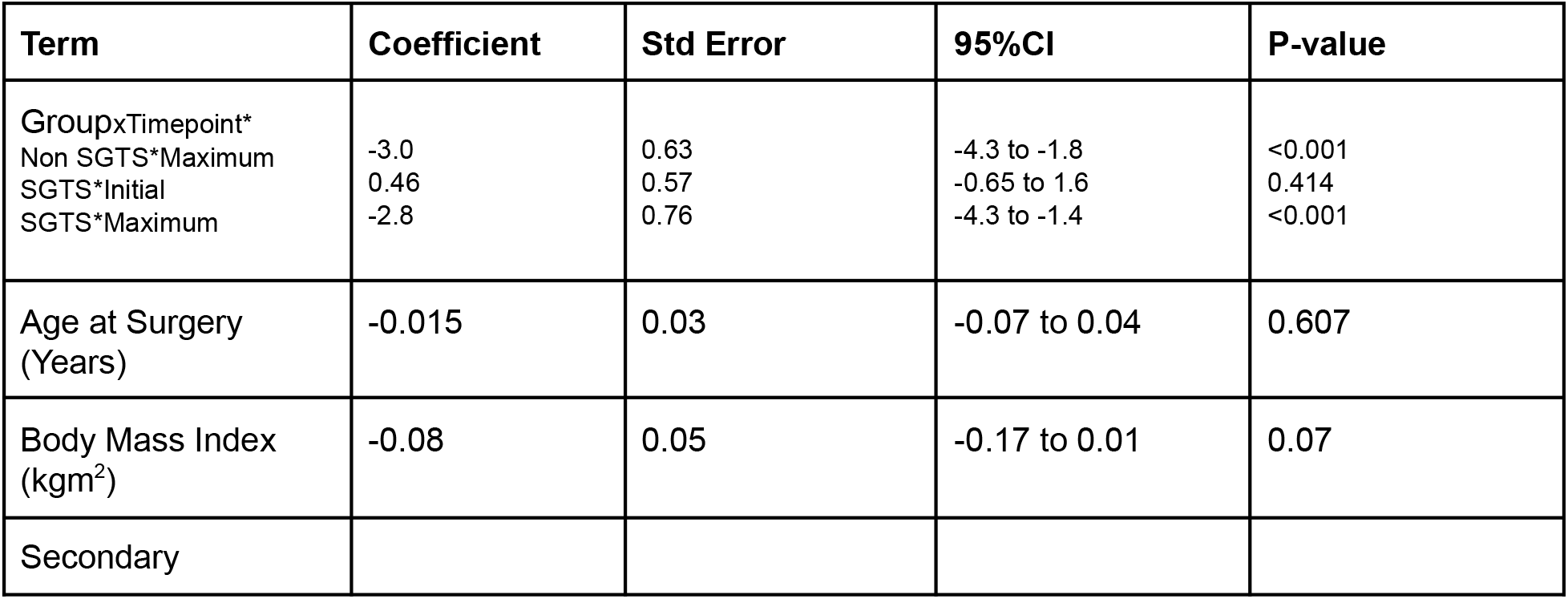
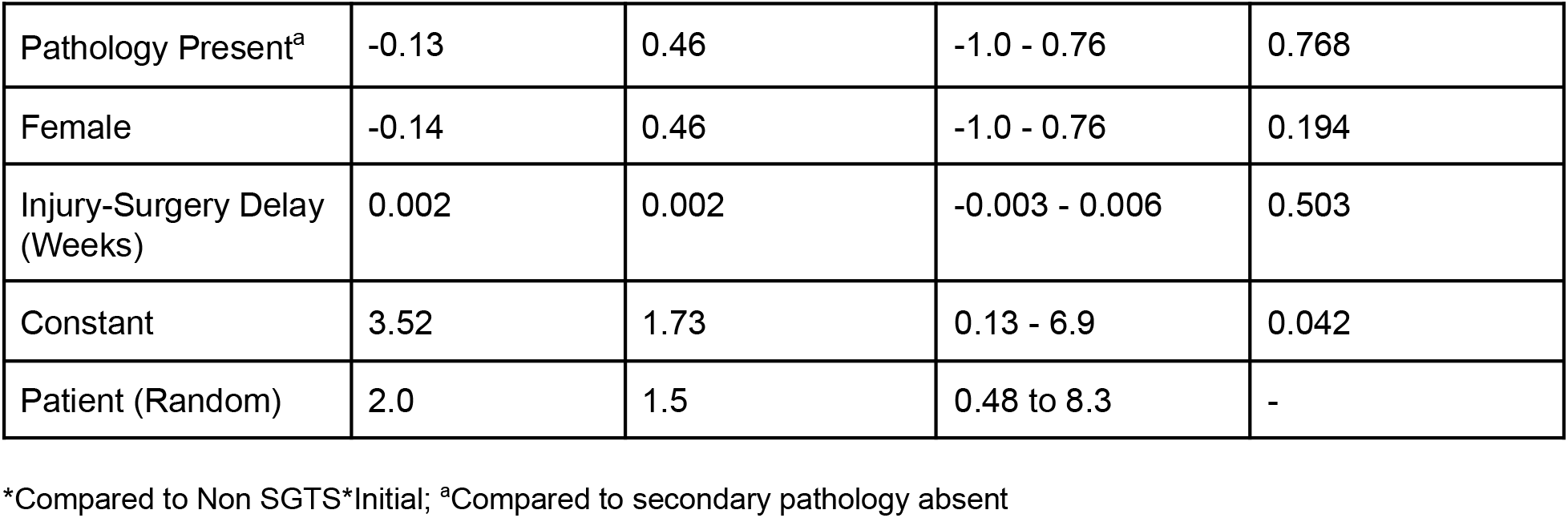
Summary of mixed-effects binary logistic regression results for LOE at Initial and Maximum Extension assessments

**Table 3:** Predicted proportion of patients presenting with LOE (95%CI) by group and time point

|  | SGTS | Non-SGTS |
| --- | --- | --- |
| Initial | 0.70 (0.56 - 0.84) | 0.62 (0.52 - 0.72) |
| Maximum | 0.17 (0.06 - 0.29) | 0.15 (0.08 - 0.23) |

## 4. Discussion

This study has demonstrated that SGTS tensioned in full extension is not inferior to other constructs for hamstring ACLR reconstruction with respect to loss of extension (LOE) measured clinically during acute, supervised rehabilitation. The upper confidence interval for the difference in LOE at both points of measurement were well below the non-inferiority margin.

Surgeons performing ACL reconstruction using hamstring autograft commonly tension the graft in 30° of flexion [16, 20]. Due to the increased diameter and stiffness of the SGTS graft compared to traditional hamstring autograft techniques, tensioning in flexion risks persistent LOE leading to inferior patient outcomes. This study demonstrated that by tensioning the SGTS graft in full extension, the patient is not at increased risk of LOE. LOE is one of the most common complications following ACL reconstruction [8, 21]. Shelbourne [27] demonstrated that loss of 5 deg of extension compared to the non-operative side was the strongest predictor of low subjective postoperative scores. In addition, persistent LOE was shown to correlate with development of osteoarthritis [18, 25, 26]. In a multicentre study comprising 3633 patients, Delaloye [9] demonstrated that early LOE was associated with reoperation for cyclops syndrome with an 8-fold increased risk if present at 6 weeks post-operatively.

LOE is associated with a decreased ability to load the operative limb during rehabilitation. Garrison [13] demonstrated that an extension deficit in rehabilitation was correlated with decreased quadriceps force resulting in increased energy expenditure on return to sport. Labanca [17] found that LOE up to two months post-reconstruction was correlated with persistent decreased knee extensor strength through to returning to sport with resultant asymmetric limb loading. This correlated with increased re-rupture rates.

Anatomic ACL reconstruction places the graft in a non-isometric position. Multiple authors have demonstrated that the graft elongates 1-3 mm when the knee is ranged from 30 deg of flexion to full extension [4, 19]. In a well designed cadaveric study, Austin demonstrated that tensioning the ACL graft at 30 deg of flexion resulted in 12-14 deg loss of extension regardless of the amount of tension applied to the graft [1].

Despite this knowledge, surgeons using hamstring autograft continue to tension the graft with the knee in flexion. In a survey of Australian orthopaedic surgeons, Kirwan reported that 40% tensioned the ACL graft in 30° of flexion [16]. This apparent lack of concern for LOE by surgeons using hamstrings for ACL reconstruction can be explained by the correlation between initial graft tension and knee stability [12]. Though not evidence-based, there is a perception that over-constraining the knee by tensioning the graft in flexion will ultimately lead to full range of motion and a more stable knee due to graft elongation. As hamstring grafts have been shown to provide less stability than bone-patella-tendon-bone grafts, this tensioning technique may be used as an attempt to rectify this deficiency [3, 11].

After creation of this shorter, thicker graft, the TLS technique recommends 500N of pre-tension be applied for 1-2 minutes. This theoretically minimises the degree of further elongation possible in vivo. There may also be less slippage between the interference screw and the tape used in the TLS technique compared to interference of a screw against the graft [2].

Impingement of the graft against the intercondylar notch is also correlated with LOE [14]. It is common to correct this impingement intra-operatively by performing a notchplasty. The rate of notchplasty was significantly higher in the non-SGTS group in this study (44.7% v 17.8%). This suggests that a higher proportion of patients in the non-SGTS group had grafts that were impinging, however, the decision to perform a notchplasty is subjective and thus this should be interpreted with caution. As the incidence of LOE in the non-SGTS group was within an acceptable range, it does not appear the higher rates of notchplasty in this group had an effect on LOE.

A short period of immobilisation was prescribed in a higher proportion of patients undergoing non-SGTS reconstructions (46.8% v 24.4%). In addition to tensioning in flexion, this may represent another technique to prevent delayed joint laxity after hamstring graft elongation. Historically, post-operative immobilisation could be prolonged and correlated with LOE [8]. The majority of prescribed movement restrictions in this study were less than two weeks, however, and were designed to protect the graft before quadriceps control was regained. This practice has not been shown to correlate with LOE, making its efficacy uncertain.

This study should be interpreted in the context of its limitations. The retrospective design increases the likelihood that recognised and unrecognised confounding factors may influence the results. Though every attempt was made to statistically control for factors known to affect LOE, this can only truly be determined with a prospective randomised controlled trial. In addition, the non-SGTS group was heterogeneous. The 12 surgeons from this group all utilised hamstring grafts tensioned at 20-30° of flexion, however there were 4 different fixation techniques utilised, each with its own biomechanical properties. Though this may improve generalisability, further work may be required to discern LOE risk with SGTS compared to a specific alternative preparation/fixation method. Follow up was relatively short (median 3 weeks) as the patients were only monitored until completion of formal rehabilitation with physiotherapy. Longer duration studies are required to determine the effects of the SGTS technique on functional outcomes and rerupture rates.

In conclusion, surgeons using or considering using the SGTS construct can rule out increased incidence of LOE as a factor in their decision-making, providing they tension the graft in full extension. Surgeons using this technique can capitalise on the theoretical benefits of a stiffer, thicker hamstring graft, without the additional donor site morbidity associated with non-hamstring grafts of similar properties such as bone-patella-tendon-bone or quadriceps tendon. Physiotherapists/allied health involved in the assessment of post-reconstruction patients are cautioned against assumptions regarding surgical technique and LOE in this population and are encouraged to explore other mechanisms of LOE causation.

## Supporting information

STROBE checklist

## Data Availability

Blinded data may be available from the authors at reasonable request.

## Acknowledgements

The authors thank Meredith Harrison-Brown for her assistance with editing and formatting the manuscript.

