## Supplementary material for "The effect of short-graft preparation with tape suspension and screw fixation on loss of knee extension following anterior cruciate ligament reconstruction: A retrospective cross-sectional analysis of public hospital cases from 2015 - 2017"

**Supplementary table:** Variables included in regression model

| Variable | Definition | Responses | Component of model |
|---|---|---|---|
| Patient ID | Unique identifier | ID, URN, MRN | Model random factor |
| Age | Age at time of surgery | years | Model covariate |
| Gender | As identified by the patient | Male; Female; Not specified | Model covariate |
| BMI | Body mass index as per standard formula | kg/(m^2) | Model covariate |
| Secondary diagnosis | Injuries to other knee structures | meniscus (lateral/medial); cartilage (femoral/tibial/patella); ligament (collateral/PCL); other | Model covariate |
| DateofInjury to DateofSurgery | Time delay between when the injury occurred and when surgery is performed | Weeks | Model covariate |
| Timepoint | Post-surgery delay of measurement of maximum knee flexion | Initial<br>Maximum | Model covariate |
| SGTS | Was a SGTS system used? | Yes; No | Model primary predictor |
| Maximum extension | Minimum flexion angle under passive loading forcing the knee into extension. Relative to anatomical zero | Degrees | Model primary outcome |

**Supplementary data:** Regression model code (STATA)

```
import delimited "Filepath MasterSheet.csv", stringcols(1)

*Swap Filepath for local filepath

// Mixed effects logistic regression as per DAG figure (adjusted variables as covariates) (updated Sept 2021)

* encode categorical variables
label define sexcode 0 "Male" 1 "Female", replace
encode sex, gen(sexcode)
encode secondary, gen(secondcode)
label define sgtscode 0 "No" 1 "Yes", replace
encode sgts, gen(sgtscode)
label define timecode 0 "Initial" 1 "Maximum", replace
encode timepoint, gen(timecode)
label define loecode 0 "No" 1 "Yes", replace
encode loestatus, gen(loecode)
label define notchcode 0 "No" 1 "Yes", replace
encode notchplasty, gen(notchcode)
encode secmed, gen(secmedcode)
encode seclat, gen (seclatcode)
encode secother, gen(othercode)
encode presromrest, gen(presromcode)

*Check missing pattern of key variables
misstable summarize, all
misstable patterns ageatsurgery sexcode secondcode dos_physio bmi sgtscode timecode

*Sort data to summarize
sort timecode

*Create table summary by timepoint
*by timecode: summarize, separator(4)
table sgtscode notchcode, statistic (count notchcode)

table sgtscode, statistic(count ageatsurgery bmi dos_physio doi_dos initial_max)
table sgtscode, statistic(median ageatsurgery bmi dos_physio doi_dos initial_max)
table sgtscode, statistic(q1 ageatsurgery bmi dos_physio doi_dos initial_max)
table sgtscode, statistic(q3 ageatsurgery bmi dos_physio doi_dos initial_max)

*Set and perform melogit

*label define loecode 0 "No" 1 "Yes", replace
melogit loecode i.sgtscode#i.timecode c.ageatsurgery c.secondcode c.bmi i.sexcode c.doi_dos, || patientid:
// Calculated proportions for sgtscode
margins sgtscode
// Calculated proportions for sgtscode
margins sgtscode#timecode
// Calculate differences in proportions between TLS levels (Yes vs No)
margins ar.sgtscode
// Calculate proportions for sgtscode##timecode
margins ar.sgtscode@timecode

save "Filepath Analysis - First Pass.dta", replace
*Swap Filepath for local filepath
```
