## Supplementary material for "The effect of short-graft preparation with tape suspension and screw fixation on loss of knee extension following anterior cruciate ligament reconstruction: A retrospective cross-sectional analysis of public hospital cases from 2015 - 2017": STROBE checklist

| | Item No | Recommendation | Yes/no | Page |
|---|---|---|---|---|
| Title and abstract | 1 | (a) Indicate the study's design with a commonly used term in the title or the abstract | Yes | 1 |
| | | (b) Provide in the abstract an informative and balanced summary of what was done and what was found | Yes | 1-2 |
| **Introduction** | | | | |
| Background/rationale | 2 | Explain the scientific background and rationale for the investigation being reported | Yes | 3-4 |
| Objectives | 3 | State specific objectives, including any prespecified hypotheses | Yes | 4 |
| **Methods** | | | | |
| Study design | 4 | Present key elements of study design early in the paper | Yes | 5 |
| Setting | 5 | Describe the setting, locations, and relevant dates, including periods of recruitment, exposure, follow-up, and data collection | Yes | 5 |
| Participants | 6 | (a)Cohort study—Give the eligibility criteria, and the sources and methods of selection of participants. Describe methods of follow-up<br>Case-control study—Give the eligibility criteria, and the sources and methods of case ascertainment and control selection. Give the rationale for the choice of cases and controls<br>Cross-sectional study—Give the eligibility criteria, and the sources and methods of selection of participants | Yes | 5-6 |
| | | (b)Cohort study—For matched studies, give matching criteria and number of exposed and unexposed<br>Case-control study—For matched studies, give matching criteria and the number of controls per case | NA | |
| Variables | 7 | Clearly define all outcomes, exposures, predictors, potential confounders, and effect modifiers. Give diagnostic criteria, if applicable | Yes | 6-8 |
| Data sources/ measurement | 8* | **For each variable of interest, give sources of data and details of methods of assessment (measurement). Describe comparability of assessment methods if there is more than one group** | Yes | 7-8 |
| Bias | 9 | Describe any efforts to address potential sources of bias | Yes | 8 |
| Study size | 10 | Explain how the study size was arrived at | Yes | 8 |
| Quantitative variables | 11 | Explain how quantitative variables were handled in the analyses. If applicable, describe which groupings were chosen and why | Yes | 9-10 |
| Statistical methods | 12 | (a) Describe all statistical methods, including those used to control for confounding | Yes | 9-10 |
| | | (b) Describe any methods used to examine subgroups and interactions | | |
| | | (c) Explain how missing data were addressed | Yes | 8-9 |
| | | (d)Cohort study—If applicable, explain how loss to follow-up was addressed<br>Case-control study—If applicable, explain how matching of cases and controls was addressed<br>Cross-sectional study—If applicable, describe analytical methods taking account of sampling strategy | Yes | 10 |
| | | (e) Describe any sensitivity analyses | NA | |
| **Results** | | | | |
| Participants | 13* | (a) Report numbers of individuals at each stage of study—eg numbers potentially eligible, examined for eligibility, confirmed eligible, included in the study, completing follow-up, and analysed | Yes | 12 |
| | | (b) Give reasons for non-participation at each stage | Yes | 12 |
| | | (c) Consider use of a flow diagram | Yes | 12 |
| Descriptive data | 14* | (a) Give characteristics of study participants (eg demographic, clinical, social) and information on exposures and potential confounders | Yes | Table 1 |
| | | (b) Indicate number of participants with missing data for each variable of interest | Yes | 13 |
| | | (c)Cohort study—Summarise follow-up time (eg, average and total amount) | Yes | 13 |
| Outcome data | 15* | *Cohort study—Report numbers of outcome events or summary measures over time* | Yes | 14 |
| | | *Case-control study—Report numbers in each exposure category, or summary measures of exposure* | NA | |
| | | *Cross-sectional study—Report numbers of outcome events or summary measures* | NA | |
| Main results | 16 | (a) Give unadjusted estimates and, if applicable, confounder-adjusted estimates and their precision (eg, 95% confidence interval). Make clear which confounders were adjusted for and why they were included | Yes | 13-14 |
| | | (b) Report category boundaries when continuous variables were categorized | NA | |
| | | (c) If relevant, consider translating estimates of relative risk into absolute risk for a meaningful time period | NA | |
| Other analyses | 17 | Report other analyses done—eg analyses of subgroups and interactions, and sensitivity analyses | NA | |
| **Discussion** | | | | |
| Key results | 18 | Summarise key results with reference to study objectives | Yes | 15-17 |
| Limitations | 19 | Discuss limitations of the study, taking into account sources of potential bias or imprecision. Discuss both direction and magnitude of any potential bias | Yes | 17 |
| Interpretation | 20 | Give a cautious overall interpretation of results considering objectives, limitations, multiplicity of analyses, results from similar studies, and other relevant evidence | Yes | 17-18 |
| Generalisability | 21 | Discuss the generalisability (external validity) of the study results | Yes | 17 |
| **Other information** | | | | |
| Funding | 22 | Give the source of funding and the role of the funders for the present study and, if applicable, for the original study on which the present article is based | NA | |
